# Brain White Matter Microstructure Associations with Blood Markers of the GSH Redox cycle in Schizophrenia

**DOI:** 10.1101/2024.09.02.24312940

**Authors:** Tommaso Pavan, Pascal Steullet, Yasser Alemán-Gómez, Raoul Jenni, Zoé Schilliger, Martine Cleusix, Luis Alameda, Kim Q. Do, Philippe Conus, Patric Hagmann, Daniella Dwir, Paul Klauser, Ileana Jelescu

**Affiliations:** Department of Radiology, Lausanne University Hospital (CHUV) and University of Lausanne (UNIL), Lausanne, Switzerland; Center for Psychiatric Neuroscience, Department of Psychiatry, Lausanne University Hospital and the University of Lausanne, Lausanne, Switzerland; Service of General Psychiatry, Treatment and Early Intervention in Psychosis Program. Lausanne University Hospital (CHUV), Lausanne, Switzerland; Department of Psychosis Studies, Institute of Psychiatry, Psychology and Neuroscience. King’s College of London, London, UK; Centro Investigacion Biomedica en Red de Salud Mental (CIBERSAM); Instituto de Biomedicina de Sevilla (IBIS), Hospital Universitario Virgen del Rocio, Departamento de Psiquiatria, Universidad de Sevilla, Sevilla, Spain; Service of Child and Adolescent Psychiatry, Department of Psychiatry, Lausanne University Hospital and the University of Lausanne, Lausanne, Switzerland

## Abstract

In groups of patients suffering from schizophrenia (SZ), redox dysregulation was reported in both peripheral fluids and brain. It has been hypothesized that such dysregulation, including alterations of the glutathione (GSH) cycle could participate in the brain white matter (WM) abnormalities in SZ due to the oligodendrocytes’ susceptibility to oxidative stress.

In this study we aim to assess the differences between 82 schizophrenia patients (PT) and 86 healthy controls (HC) in GSH-redox peripheral blood markers: GSH peroxidase (GPx), reductase (GR) enzymatic activities and their ratio (GPx/GR-ratio), evaluating the hypotheses that alterations in the homeostasis of the systemic GSH cycle may be associated with pathological mechanisms in the brain WM in PT. To do so, we employ the advanced diffusion MRI methods: Diffusion Kurtosis Imaging (DKI) and White Matter Tract Integrity-Watson (WMTI-W), which provide excellent sensitivity to demyelination and neuroinflammation.

We show that GPx levels are higher (p=0.00041) in female control participants and decrease with aging (p=0.026). We find differences between PT and HC in the association of GR and mean kurtosis (MK, p<0.0001). Namely, lower MK was associated with higher blood GR activity in HC, but not in PT, suggesting that high GR activity (a hallmark of reductive stress) in HC was linked to changes in myelin integrity. However, GSH-redox peripheral blood markers did not explain the WM anomalies detected in PT, or the design of the present study could not detect subtle phenomenon, if present.

## Introduction

Psychosis and schizophrenia (SZ) present significant challenges for patients, their families, and society (Wittchen et al., 2011). Only partially understood, its etiology is attributed to a multifaceted interplay of genetic, environmental and developmental factors that affect a variety of biological processes, including the redox balance (McCutcheon et al., 2020; Perkins et al., 2020). In SZ, a redox dysregulation was reported in peripheral fluids, suggested by a diminished antioxidant capacity, oxidative damage of lipids, DNA, and RNA (Jorgensen et al., 2022; Rambaud et al., 2022; Tsugawa et al., 2019), while in the brain, lower glutathione (GSH), and lower NAD+/NADH ratio were found (Das et al., 2019; Kim et al., 2017). The causes of such redox dysregulation and the resulting oxidative stress are multifactorial, including, chronic subclinical inflammation and weakened antioxidant capacities. Anomalies in the GSH redox system, a key antioxidant system for cell protection and detoxification, have been widely observed in SZ. Meta-analyses showed lower mean levels of GSH in blood (Tsugawa et al., 2019), and brain (Das et al., 2019; Gawryluk et al., 2011; Yao et al., 2006; Zhang et al., 2018) in patients vs. controls, as well as higher variability of brain GSH levels in SZ patients than in healthy subjects, suggesting the presence of subgroups of patients with different regulation of the GSH system (Palaniyappan et al., 2021).

The redox regulation and antioxidant function of GSH acts largely via the GSH redox cycle that involves the GSH peroxidase (GPx) and GSH reductase (GR). GPx neutralizes peroxides by oxidizing GSH into GSH-disulfide (GSSG). In turn, GR reduces GSSG back to GSH. The ratio between GSSG and GSH determines the redox potential of the GSH system that plays a central role in regulating redox-sensitive signaling pathways and mitochondrial oxidative phosphorylation. Low redox potentials (caused by too low GSH/GSSG ratio) and too high redox potentials (caused by too high GSH/GSSG ratio) are both detrimental to proper cellular function by provoking respectively oxidative stress and reductive stress. Reductive stress associated with too negative redox potential of the GSH/GSSG couple can indeed trigger mitochondrial oxidation and reduce cell viability (Huali Zhang et al., 2012). A proper coordination of GPx and GR activities is therefore essential to neutralize reactive oxygen species and maintain a proper redox potential of the GSH/GSSG couple.

Hence, GPx and GR activities and the GPx/GR-ratio could inform about the homeostasis of the GSH redox cycle. Two out of three postmortem studies have found lower brain GPx in SZ patients vs. controls (Gawryluk et al., 2011; Yao et al., 2006; Zhang et al., 2018). Similarly, meta-analyses suggest that SZ patients tend to also display lower blood GPx than controls (Tsugawa et al., 2019). However, the decrease in peripheral GPx activity seems to be mostly found in chronic patients (Flatow et al., 2013). In contrast to GPx, there is no evidence of consistent alterations of GR in the brain (Yao et al., 2006) or the blood (Tsugawa et al., 2019).

A possible reason for the discrepancies between studies is the influence of confounding factors such as age (Jones et al., 2002; Martínez de Toda et al., 2019), sex (Schilliger et al., 2024) and possibly medication (Raffa et al., 2009). This highlights the importance of accounting for the confounding effects of these demographic variables on the GSH-redox measures in relation to disease.

It has been hypothesized that a GSH deficit could contribute to the brain white matter (WM) abnormalities in SZ (Monin et al., 2016, Monin et al., 2015), due to the susceptibility of oligodendrocytes to oxidative stress (Juurlink et al., 1998) as a consequence of their elevated metabolic activity and iron content required for myelin production and maintenance (Bradl and Lassmann, 2010; Thorburne and Juurlink, 1996).

The GSH redox cycle supports the WM integrity by promoting the survival, proliferation and differentiation of the oligodendrocyte precursor cells (Back et al., 1998; Monin et al., 2015), protecting the oligodendrocytes from oxidative stress thanks to the GPx expression (Baud et al., 2004; Ravera et al., 2015). Beyond oligodendrocytes, the GSH system dysregulation can also impair the energy metabolism of axons (Nave et al., 2023), the mitochondrial functioning (Smith et al., 2019), and lead to subclinical activation of microglia (Dwir et al., 2020), affecting the WM. Post-mortem studies of SZ patients have reported reduced oligodendrocyte densities in the cortex and subcortical regions (Bernstein et al., 2009; Hof et al., 2003), swollen or dystrophic oligodendroglia (Uranova et al., 2018, Uranova et al., 2011; Williams, 2021), adjacent microglia (Uranova et al., 2020), and decompaction and splitting of the myelin sheath, with vacuole inclusions between the myelin sheath (Uranova et al., 2018, Uranova et al., 2011; Williams, 2021).

Abnormal white matter integrity in SZ is also widely reported using *in-vivo* diffusion magnetic resonance imaging (dMRI). Most studies investigating schizophrenia show widespread increases in mean diffusivity (MD) and/or reductions in fractional anisotropy (FA) (Barth et al., 2023; Cetin-Karayumak et al., 2020; Kelly et al., 2018) with medium effect sizes (Kelly et al., 2018) (*Cohen’s d*=0.42), consistent with anomalies observed in the *post-mortem* studies.

However, direct evidence linking white matter abnormalities to the GSH system in schizophrenia remains limited. GSH levels in frontal brain regions have been found to correlate positively with generalized FA in the cingulum of both early psychosis patients and controls (Monin et al., 2015), while it was shown in earlier work that the precursor of GSH and antioxidant, N-acetylcysteine, improves the integrity of the fornix (i.e., increases generalized FA) (Klauser et al., 2018) and increases brain GSH (Conus et al., 2018) in early psychosis patients.

Here, we aim to assess whether the activity of GPx, GR, and the GPx/GR ratio measured in blood cells (which inform about the systemic homeostasis of the GSH redox cycle) are associated with the brain WM microstructure estimated using dMRI, in patients with a diagnosis from the schizophrenia spectrum. For this purpose, we utilized advanced dMRI methods: Diffusion Kurtosis Imaging (DKI) (Jensen et al., 2005; Jensen and Helpern, 2010), an extension of diffusion tensor imaging (DTI), and the biophysical model White Matter Tract Integrity-Watson (Fieremans et al., 2011; Jespersen et al., 2018) (WMTI-W). In addition to classical DTI measures such as MD and FA, DKI enables the estimation of the mean kurtosis (MK), a measure that provides complementary information about tissue heterogeneity. The WMTI-W model allows the estimation of specific microstructural cellular features such as the axonal density (*f*) in each voxel with fewer *ad hoc* simplifying assumptions and fit constraints (Novikov et al., 2018; Hui Zhang et al., 2012), and increased validity (Novikov et al., 2018) as compared to other biophysical methods such as free-water imaging (Pasternak et al., 2009) (FWI) and Neurite Orientation Dispersion and Density Imaging (NODDI) (Hui Zhang et al., 2012). We chose WMTI-W over FWI, which is commonly found in studies on inflammation in schizophrenia, due to reserves about interpreting the free-water fraction as a direct marker of inflammation. Notably, diffusivities tend to decrease during acute gliosis, challenging the specificity of free-water to inflammation (Guglielmetti et al., 2016). The FWI model also separates signal contributions into free isotropic water and an anisotropic tissue compartment, which conflates intra- and extracellular components. In contrast, our use of the Standard Model of white matter (like WMTI-W) provides greater specificity in distinguishing inflammation from cell loss or myelin damage (Jelescu and Fieremans, 2023).

We focused our analysis on these four metrics (MD, FA, MK, *f*) because of several factors. Firstly, we wanted to limit the number of multiple comparisons to retain good statistical power. Furthermore, MD, FA and MK are complementary and highly sensitive to changes in the tissue microstructure, with similar or higher effect size as compared to RD and RK in early psychosis and schizophrenia (Pavan et al., 2025). As for WMTI-W metrics, *f* was shown to be sensitive to both demyelination and changes in axonal densities at earlier stages than the extra-axonal perpendicular diffusivity (Falangola et al., 2014; Jelescu et al., 2016), and showed similar sensitivity to the extra-axonal perpendicular diffusivity in discriminating groups (Pavan et al., 2025). Together, this set of metrics has the ability to detect abnormal myelination or axonal densities (⍰MD, ⍰FA, ⍰MK, ⍰*f*) (Falangola et al., 2014; Guglielmetti et al., 2016; Jelescu et al., 2016; Wang et al., 2011), but also neuroinflammation which has the effect of reducing diffusivities (⍰MD) and increasing kurtosis and apparent axonal water fraction (⍰MK, ⍰*f*) due to higher cellular crowding associated with microgliosis and astrocytosis (Guglielmetti et al., 2016; Jelescu et al., 2016; Jelescu and Fieremans, 2023; Wijtenburg and Rowland, 2023).

In our previous work using this same cohort, which focused on WM alterations in EP and SZ vs healthy controls (HC) (Pavan et al., 2025). We showed that WM MD was higher, while FA, MK, and *f* were lower in early psychosis and SZ (⍰MD, ⍰FA, ⍰MK, ⍰*f*) than in HC, suggesting the predominance of myelin integrity abnormalities rather than active inflammatory processes, which is also consistent with the cohort treatment history.

Here we hypothesize that alterations in the homeostasis of the systemic GSH redox cycle may be associated with pathological mechanisms in the brain (i.e., redox dysregulation and microglia dysfunction, altered oligodendrocyte density and myelination) that affect WM. Therefore, we expect that peripheral markers related to the GSH redox cycle may correlate with WM microstructure metrics sensitive to axonal density, neuroinflammation and myelination.

More specifically, we hypothesize that individuals with a high GPx/GR-ratio would exhibit a more oxidized state of the GSH/GSSG couple compared to those with a low GPx/GR-ratio. An elevated GPx/GR ratio and/or increased GPx activity in the blood may indicate systemic oxidative stress, whereas a low ratio and/or high GR activity may suggest systemic reductive stress. Notably, patients who benefited from N-acetylcysteine treatment were primarily those with high blood GPx activity (Conus et al., 2018). Conversely, reductive stress can disrupt redox-sensitive processes and paradoxically elevate mitochondrial reactive oxygen species production, ultimately impairing mitochondrial function and causing oxidative damage (Korge et al., 2015). Based on this, we stratified participants into high and low GPx/GR-ratio groups to investigate potential differences in brain microstructure related to distinct oxidative or reductive states. A comparable approach was used in a study assessing baseline GPx activity (Conus et al., 2018).

Finally, due to the known effect of age (Martínez de Toda et al., 2019; Stadtman, 2002) and sex (Martínez de Toda et al., 2023; Ruszkiewicz et al., 2019) on the expressions of the GSH redox cycle markers, we explored the role that these confounding variables could have on the link between our peripheral GSH-redox measurements and WM microstructure.

## Methods

### Participants

Data were collected from 168 individuals (**Table 1**) divided into two groups: 86 healthy controls (HC) and 82 patients (PT) with a diagnosis from the schizophrenia spectrum disorder (**Supplementary Table S1**). Patients were recruited from the Lausanne University Hospital. PT with psychosis related to intoxication or organic brain disease, IQ*<*70, reporting alcoholism, history of drug abuse (nicotine excluded), major somatic disease, documented anamnestic or current organic brain damage were excluded. HC were recruited from the same sociodemographic area as the patients. HC were excluded if they or a first-degree family member reported to have suffered from neurological, traumatic, major mood disorder, psychosis, prodromal symptoms, and current or past antipsychotic treatments. Symptoms in PT were assessed via the Positive and Negative Syndrome Scale (PANSS) (Kay et al., 1987), and global functioning was estimated for the whole sample via the Global Assessment of Function scale (GAF) (American Psychiatric Association, 2000) by a trained psychologist (not blinded). The study was approved by the local Ethics Committee of the Canton of Vaud (Switzerland) under authorization numbers CER-VD 382/11 and 2018-01731.

**Table 1.**
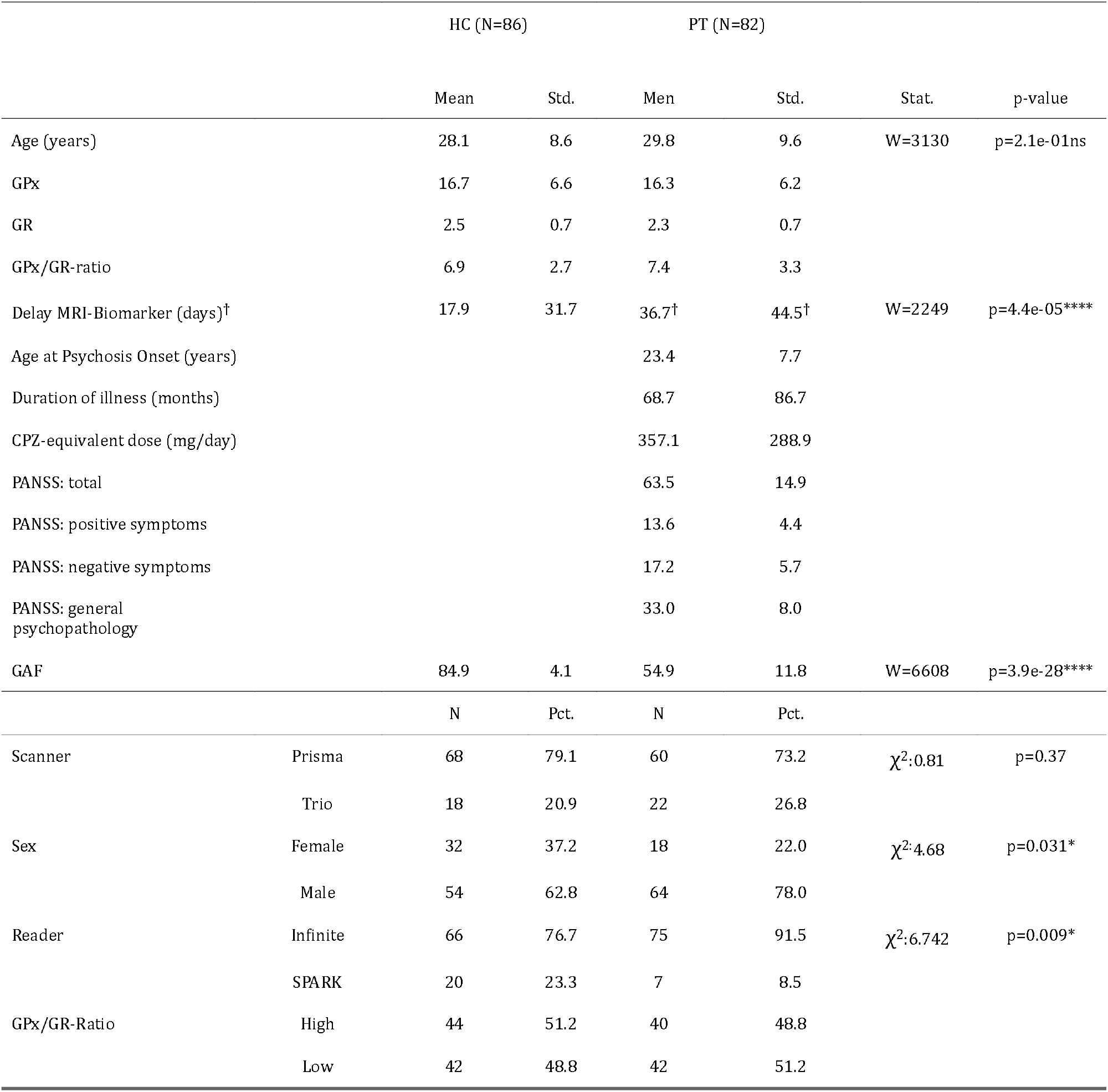
Cohort demographics. P-values refer to Wilcox’s tests between patients and healthy control. χ2 test is computed for the scanner, sex, and reader contingency tables. GPx and GR activities are expressed in nmoles NADPH used/min/g hemoglobin. HC: healthy controls, PT: patients, GPx: glutathione peroxidase, GR: glutathione reductase, GAF: Global Assessment of Functioning, PANSS: Positive and Negative Syndrome Scale. †: the delay MRI-biomarker statistics are reported excluding one PT participant with an extreme delay of 271 days.

### MRI acquisition

MRI scanning sessions were performed on two different 3-Tesla systems (Magnetom TrioTim and PRISMA, Siemens Healthineers, Erlangen, Germany), each equipped with a 32-channel head coil. A 1-mm isotropic T1-weighted image was acquired for anatomical reference. Whole-brain diffusion-weighted images (DWI) were acquired using diffusion spectrum imaging (DSI) scheme across 15 b-values, ranging from 0 to 8000 s/mm^2^, spatial resolution of 2.2 × 2.2 × 3 mm^3^ - See Supplementary Material for further acquisition details.

### Image preprocessing

The MPRAGE image was bias field corrected (Tustison et al., 2010) and skull-stripped via nonlinear registration to the MNI-152 template (Grabner et al., 2006) using Advanced Normalization Tools (ANTs) (Avants et al., 2008). The diffusion preprocessing pipeline included MP-PCA denoising, Gibbs ringing-, EPI-, eddy current and motion corrections, following most recent guidelines (Ades-Aron et al., 2018) - see Supplementary Material for preprocessing details.

### Microstructure estimation

For Diffusion Kurtosis Imaging and White Matter Tract Integrity - Watson estimation, the diffusion dataset was truncated (Jensen et al., 2005) at b≤2500 s/mm^2^ (refer to Supplementary Methods for details). DKI was fit voxel-wise in the entire brain (Veraart et al., 2013) using a weighted linear-least squares algorithm in Matlab (Veraart et al., 2013), from which seven scalar maps were derived. However, only three parameter maps were included in the statistical analysis: MD, FA, and MK. WMTI-W parameters were estimated voxel-wise from the seven DKI scalars, using an in-house Python script, yielding five parameter maps. However, only axonal density *f* was included in the statistical analysis. We focused our analysis on these four metrics (MD, FA, MK, *f*) because of their ability at detecting abnormal myelination or axonal densities (⍰MD, ⍰FA, ⍰MK, ⍰*f*) (Falangola et al., 2014; Guglielmetti et al., 2016; Jelescu et al., 2016; Wang et al., 2011), and neuroinflammation (⍰MD, ⍰MK, ⍰*f*) (Guglielmetti et al., 2016; Jelescu et al., 2016; Jelescu and Fieremans, 2023; Wijtenburg and Rowland, 2023).

### Blood analyses

Given that diurnal fluctuations are known for both GPx and GR (Budkowska et al., 2022), blood sampling was tipically done early in the morning (8-10am) under fasting conditions. GR and GPx activity were assessed in hemolyzed blood cells incubated in phosphate buffer solution (100mM, pH 7.5) containing EDTA (0.6mM), and non-limiting levels of either oxidized glutathione (GSSG, 2.5mM) and NADPH (0.25mM) (for GR), or GSH (2.5mM), NADPH (0.25mM), GR (0.84U/ml; Fluka) and tert-butyl hydroperoxide (TBHP, 0.8mM, Fluka) (for GPx). We expressed GPx and GR activities in nmoles NADPH used/min/g hemoglobin. These enzymatic activities were measured with two different plate readers. Batch effects between plate readers were corrected via ComBat for biological data (Johnson et al., 2007) (see additional details on the blood analyses and harmonization procedure in the Supplementary Methods).

### Statistical analysis

#### Blood markers analysis

Statistical analyses were run in R, group comparisons were computed via Welch’s t-test. Robust linear regression (Maechler et al., 2023) was used for all the regression and contrast analyses, always including a correction by sex and quadratic age (see Age Correction section in Supplementary Methods for additional details).

#### Association between blood markers and WM microstructure

Participants with a delay between MRI scan and blood samples larger than 60 days were excluded whenever an association between blood biomarkers and WM microstructure metrics was tested (86% of the original cohort, N_HC_=80, N_PT_=65, delay_HC_=10.0±11.7, delay_PT_=17.4±16.0, p=0.0012). The 60-days cutoff was chosen to retain the highest number of subjects while reducing the group-level delay and controlling the effect of seasonal or longer timescale fluctuations on the blood markers (see **Fig. S4** for details). All analyses included a correction for quadratic age, sex, and delay. Associations were tested at voxel level using FSL’s Tract-Based Spatial Statistics (TBSS) (Smith et al., 2006). First, individual FA maps were used to build a study-specific FA template using ANTs (Avants et al., 2008) and the estimated warps were used to spatially normalize each individual FA, MD, MK and *f* map. Before any statistical analysis, all the microstructure parameter skeletons were harmonized for scanner type at voxel level in an unbiased way (mean normalized towards the center of both sites, without reference batch) via ComBat harmonization (Fortin et al., 2017). ComBat was previously proven efficient at correcting scanner effects in the ENIGMA-consortium (Radua et al., 2020), in the current cohort (Alemán-Gómez et al., 2020; Pavan et al., 2025), and validated in travelling participants (Richter et al., 2022). After permutation testing (FSL randomize (Winkler et al., 2014), 5000 permutations), the resulting statistical maps were false discovery rate and Threshold-Free Cluster Enhancement corrected.

#### Low and High GPx/GR ratio analysis

To study if there could be differences between participants showing different levels of GPx and GR activity, we repeated the analysis described above after re-classifying the participants into two groups according to their GPx/GR-ratio. Low-ratio and high-ratio were defined as having a GPx/GR-ratio respectively lower and higher than the median GPx/GR ratio of the HC group (**Fig.1A**, Median_HC_=6.64).

**Figure 1:**
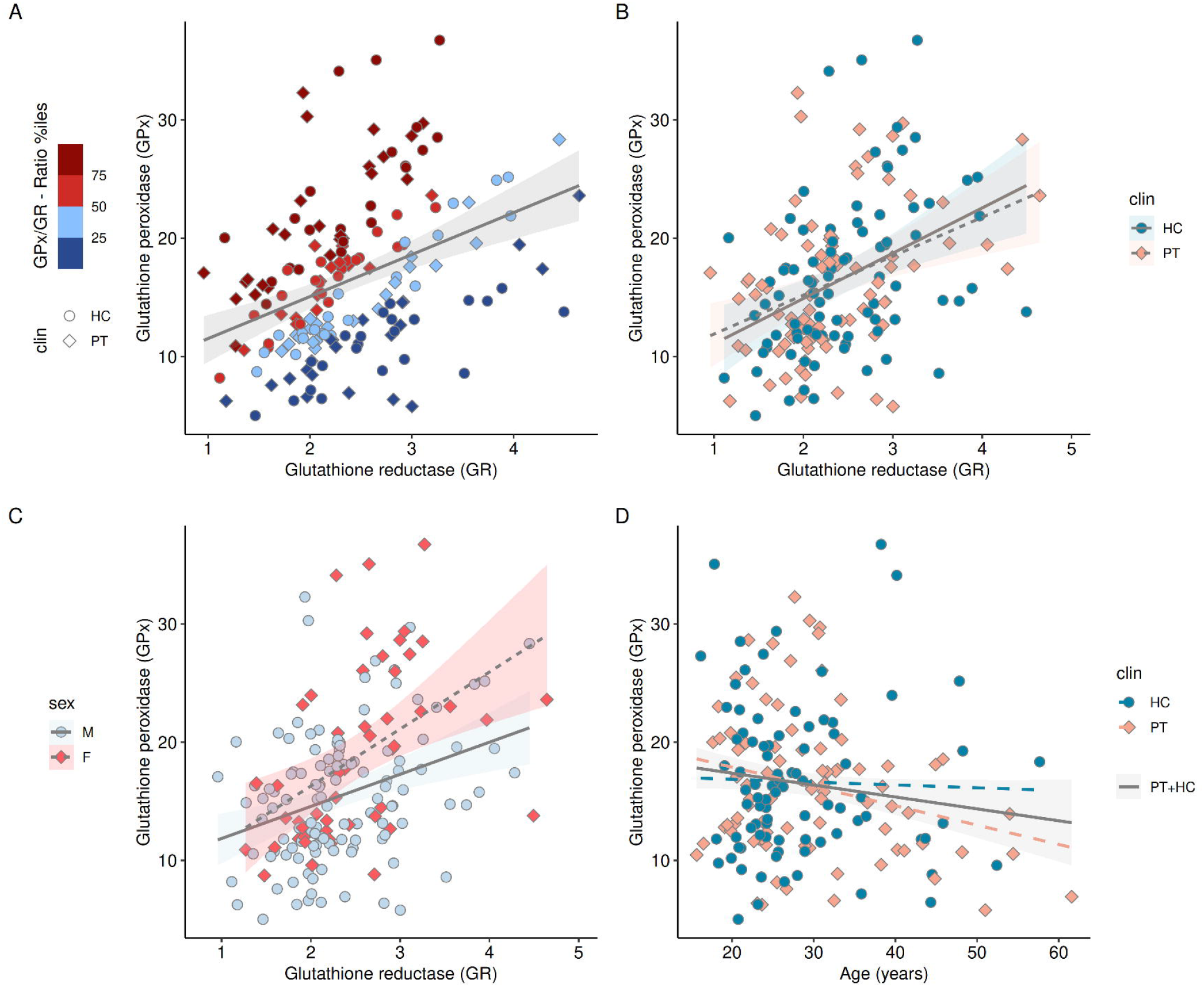
Biomarkers of the blood GSH-redox cycle and the effects of sex, age and clinical status. Correlations between GPx and GR activity in the whole cohort, colored by the quartiles of the ratio between GPx and GR (A), clinical status (B), and sex (C). Association between GPx and age (D) for both the entire cohort (shaded gray area) and by clinical status. GPx and GR activities are expressed in nmoles NADPH used/min/g hemoglobin. GPx: GSH peroxidase; GR: GSH reductase; HC: healthy controls; PT: patients; M: males; F: females.

## Results

### Demographics

Summary demographics of the cohort can be found in **Table 1**. PT (29±9 y/o) and HC (28±8 y/o) did not differ in age (p=0.21). As expected, differences were found between PT and HC in functional levels as measured by the GAF scale (p<0.0001). Differences between groups were also found in the delay between MRI acquisition and blood collection (p<0.0001), leading to the inclusion of the delay as a covariate.

### GSH-redox cycle system

Overall, GPx (p=0.69) and GR (p=0.24) activities or the GPx/GR-ratio (p=0.32) did not differ between PT and HC groups. HC female participants showed higher GPx activity than males (p=0.00041) but the same trend was not found in PT. GPx and GR activities correlated significantly for the whole cohort (r=0.38, p<0.0001, **Fig. 1A**), but also for PT (r=0.38, p=0.00038) and HC (r=0.40, p=0.00013) when considered independently. The slope contrast between PT and HC did not show any significant difference (GPx ⍰ group interaction: p=0.80, **Fig. 1B**). GPx and GR activities correlated also in males and females separately (r_male_=0.33, p=0.00019 and r_female_=0.46, p=0.00066) with no significant sex difference for the slopes either (sex ⍰ group interaction: p=0.19, **Fig. 1C**). Age was only found significantly associated with a decrease in GPx activity when pooling HC and PT together (**Fig. 1D**, p=0.026). No significant association was found with medication. No significant association was found between psychiatric functioning/symptoms scores and redox markers after multiple comparison correction (**Table S2**).

### Associations between GSH-redox cycle and WM microstructure

TBSS analysis showed significant associations only between GR and MK in HC. No other association for PT or HC was found between GPx, GR or GPx/GR-ratio with either MD, FA, MK or *f*. In HC, MK was significantly and negatively associated with GR in multiple WM clusters (**Fig. 2A**, p=0.00018). Furthermore, the slope (GR–MK) in HC differed significantly from the regression slope of PT (**Fig. 2B**, p<0.0001). The significant contrast was localized bilaterally in the anterior and superior corona radiata, genu and body of the corpus callosum.

**Figure 2:**
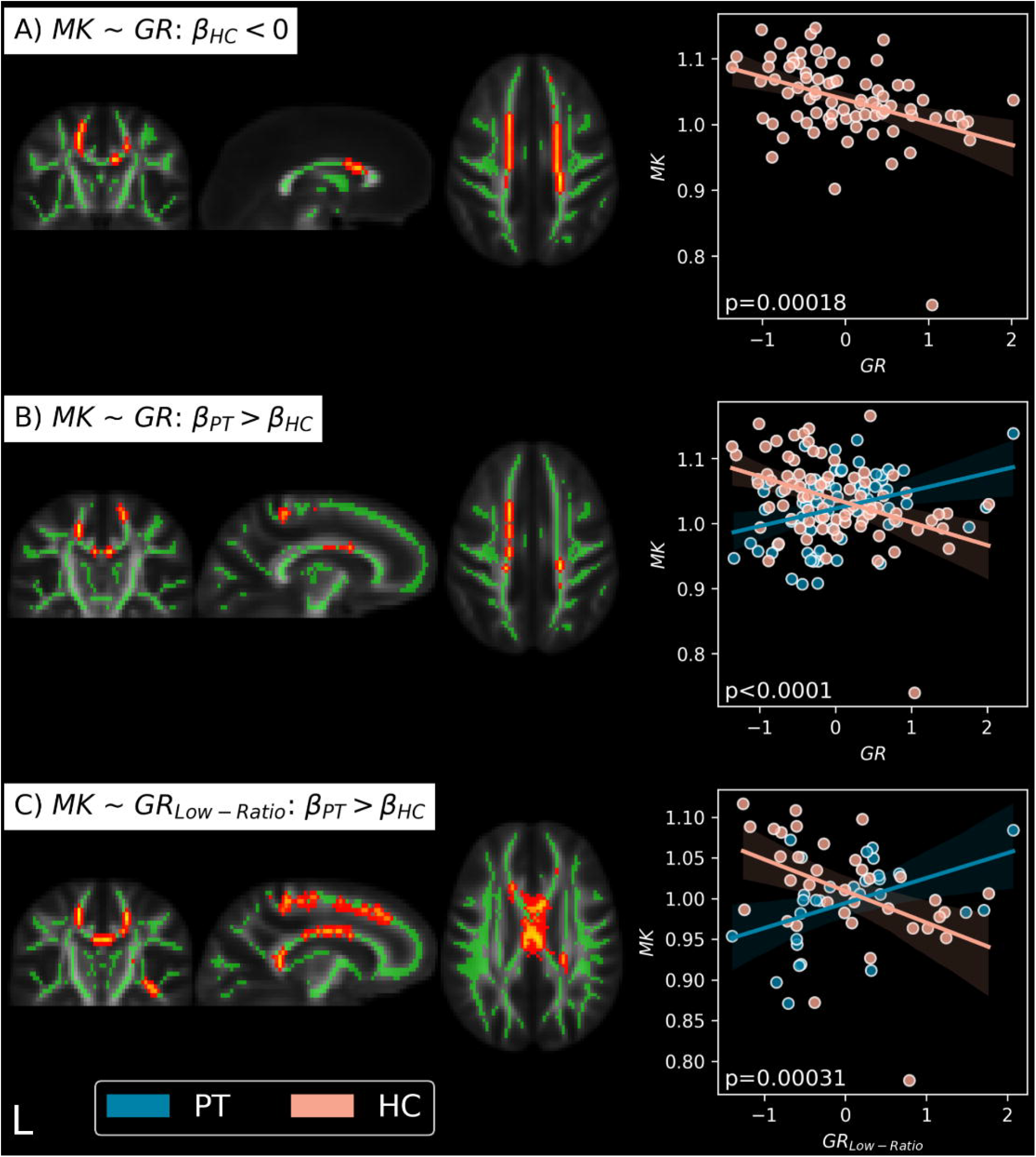
Significant associations between biomarkers of the blood GSH-redox cycle and the dMRI microstructure estimates. (A, B): Brain plots show the significant association clusters between MK and GR activity (p_fdr_≤0.05). GR was found significantly associated with MK in HC (A), with a slope that differed from the PT slope (B). (C): Brain plots show the significant clusters where the association of MK with GR in individuals with low GPx/GR-ratio yielded a different slope between HC and PT (p_fdr_≤0.05). The regression plots show the association average values within significant clusters. Clusters were identified in the bilateral anterior and superior corona radiata, as well as in the genu and body of the corpus callosum and sections of the internal capsule. Results are corrected for quadratic age, sex, and delay. GR and GR_Low-ratio_ are centered. Excluding the HC participant with MK < 0.80 does not change the results. Green: WM skeleton; HC: healthy controls; PT: patients; GR: glutathione reductase; MK: mean kurtosis; β_HC_: slope of HC; β_PT_: slope of the PT; Low-ratio: participants with GPx/GR-ratio < median ratio of the HC group (6.64).

### Associations between GSH-redox cycle and dMRI WM microstructure in the low and high GPx/GR-ratio groups

We then assessed how low and high GPx/GR-ratio reflected at the WM microstructure level. First, we established how the low-vs high-ratio is driven by both GPx and GR levels. Considering both PT and HC together, participants with low ratio (p<0.0001) had significantly lower GPx (mean GPx=13.09) and higher GR (mean GR=2.61) as compared to subjects with high ratio (mean GPx=19.86, mean GR=2.18). GPx and GR significantly correlated within both low-(r=0.75, p<0.0001) and high-ratio (r=0.67, p<0.0001) groups.

TBSS voxel-wise analysis of the 3-way interaction model of GPx or GR interacting with low/high-ratio, and clinical status, showed widespread significant differences in the slopes of MK vs GR between low ratio HC and PT (**Fig. 2C**, p=0.00031) but not for any other comparison, indicating the low-ratio group drives the difference between HC and PT found in the previous section. Identified areas for the GR-MK association were the bilateral anterior and superior corona radiata, genu, splenium and body of the corpus callosum, right posterior limb, and right retrolenticular part of the internal capsule. Notably, many voxels of the peripheral WM were also significant.

## Discussion

In this work, we quantified peripheral biomarkers of the GSH redox cycle (GPx, GR activities and their ratio) in patients with schizophrenia and control subjects, and we explored the relationships of these redox markers with the WM microstructure estimated with advanced dMRI methods. We did not find significant differences in GPx, GR or GPx/GR-ratio between groups. However, female participants showed higher GPx activity than males, independently of their clinical status. GPx was also the only biomarker whose activity decreased with age, suggesting a decrease in antioxidant defense. Finally, GR activity was negatively associated with MK in HC, but not in PT, mostly driven by the subgroup of individuals with a low GPx/GR-ratio where the differences between HC and PT were enhanced and more widespread across the WM.

### Peripheral GSH redox cycle

Many studies have investigated blood GPx and GR activities in cohorts of patients suffering from schizophrenia (Flatow et al., 2013; Koga et al., 2016; Tsugawa et al., 2019; Zhang et al., 2018), sometimes with mixed results. However, meta-analyses (Flatow et al., 2013; Tsugawa et al., 2019) show overall significantly lower GPx, but not GR, in PT as compared to HC. The analysis of Flatow et al. (2013) indicates that a decrease in GPx activity was prevalent in patients with chronic SZ or in PT with acute relapses but not with first psychosis episode.

Some of the discrepancies in findings related to GSH/GPx/GR levels among studies on PT with schizophrenia could be explained not only by the stage of the disease, but also by effects of age (Jones et al., 2002; Martínez de Toda et al., 2019; Stadtman, 2002), sex (Schilliger et al., 2024), and medication (Raffa et al., 2009) on the peripheral markers. We observed higher GPx activity in females than males in HC, and decreased GPx activity with age, independently of the clinical status. However, in our cohort, which includes PT with early psychosis and chronic SZ, we did not find any difference between PT and HC regarding GPx, GR and their ratio, even after correction for age and sex.

Other sources of variability include methodological differences in sample collection, processing, storage, and quantification (see Supplementary material in Perkins et al. (2020)).

Our findings of sex differences in HC and age effects on blood GPx activity regardless of the clinical status, corroborate results from numerous studies (Abdalla et al., 1986; Jones et al., 2002; Martínez de Toda et al., 2019; Schilliger et al., 2024; Stadtman, 2002). Higher levels of GPx, but not GR, have been widely reported in the brain of females as compared to males in both animal and human studies (Dukhande et al., 2009; Martínez de Toda et al., 2023; Ruszkiewicz et al., 2019). A higher GPx activity in females is consistent with more efficient antioxidant systems and less basal inflammation in females than in males (Martínez de Toda et al., 2023). Our observation of a diminished GPx activity with age is also in line with the literature (Martínez De Toda et al., 2019) and with the notion that the antioxidant defenses weaken over the years, contributing to the accumulation of oxidative stress and its harmful effects (Stadtman, 2002).

In our work, we found that GPx and GR activities correlated positively within both PT and HC and within both males and females, suggesting a generally quite well-coordinated GSH redox cycle. Likewise, the GPx/GR ratio, which was not yet examined before to our knowledge, did not differ between PT and HC groups. Previous work showed that the activities of both enzymes were correlated in female but not male adolescents from the general population (Schilliger et al., 2024), while, EP patients with a history of childhood trauma exhibited impaired coordination between GPx and GR activity (Alameda et al., 2018). This suggests that the relationship between GPx and GR activities may differ in a sex-dependent manner during adolescence but also among subgroups of individuals according to environmental and other yet unidentified factors.

### Associations between peripheral GSH redox cycle and WM microstructure

Due to the diversity in the GSH redox cycle status across individuals and the complex effects of sex and age, it was therefore essential to tackle the relationships between the peripheral redox markers and WM microstructure by carefully considering age, sex and their interactions, and by investigating the links between the GSH redox cycle and WM separately in subgroups of individuals characterized by different peripheral redox profiles (oxidative vs reductive stress).

The strongest differences between PT and HC were found in the association of GR with the WM microstructure (**Fig. 2B, C**), particularly with mean kurtosis. MK is considered a measure of tissue complexity (Jensen et al., 2005; Jensen and Helpern, 2010), which has demonstrated sensitivity at detecting abnormal myelination (Falangola et al., 2014; Guglielmetti et al., 2016; Jelescu et al., 2016; Wang et al., 2011), neurodegeneration and neuroinflammation (Jelescu and Fieremans, 2023). Our previous work on the same data focusing on JHU region-of-interest (ROI) analysis demonstrated that MK was significantly lower in early psychosis and chronic SZ as compared to age-range matched controls (Pavan et al., 2025). Our current voxel-wise analysis on the white matter skeleton revealed that lower MK was associated with higher blood GR activity in HC, but not in patients. This suggests that high GR activity was linked with changes in myelin and tissue heterogeneity in HC. As high GR activity may be a sign of reductive stress, we looked at the relationship between GR and MK in individuals with low GPx/GR-ratio, which proved to be much stronger in this sub-group. This confirmed that the difference in the GR-MK association between PT and HC was indeed mostly driven by individuals displaying a low GPx/GR-ratio.

Furthermore, it suggests that reductive stress in blood is linked to a low MK in the white matter of healthy volunteers. Under the assumption that reductive stress in blood is accompanied by similar reductive stress in the brain, our data would indicate that white matter is susceptible to reductive stress in the form of reduced complexity, possibly from demyelination and cell density reduction. The latter could be a consequence of impaired metabolism regulation, inflammation and cytotoxicity that are known to be associated with reductive stress (Huali Zhang et al., 2012). However, high GR is not associated with low MK in the white matter of patients. The lack of a direct association between MK and GR in patients suggests that other competing mechanisms contribute more to the WM integrity impairment in patients.

Prior research has shown that N-acetylcysteine treatment can improve brain GSH levels after six months (Conus et al., 2018) and WM integrity as measured by generalized FA in PT (Klauser et al., 2018). Notably, improvements in cognitive abilities and in symptoms (Conus et al., 2018), particularly in individuals with high GPx levels, have also been reported, highlighting the interplay between blood markers, WM integrity and cognitive function. Here, however, we found no association of GPx or high GPx/GR-ratio with WM microstructure in PT, contrary to our expectation. Instead, our results showed that PT and HC had a similar redox profile with no significant difference in blood markers, indicating that patients do not exhibit a redox imbalance at the group level, which could explain the lack of correlations between blood markers and WM alterations. No patient sub-group or cluster that would show a significant difference in redox balance to the HC could be identified either.

Furthermore, Lesh et al. (2021) correlated average free-water fraction of the WM and GM with magnetic resonance spectroscopy measures of GSH/creatine ratio in the dorsolateral prefrontal cortex. The authors found a negative association between GSH/creatine and the free-water fraction in the GM but not WM, possibly suggesting that changes due to GSH, and GPx/GR, may be more evident in the GM than the WM.

Interestingly, the spatial localization of the strongest GR-MK significant contrasts between HC and patients (**Fig. 2D**) coincides with the FA WM region of interests (anterior and superior corona radiata, genu and body of the corpus callosum) showing the strongest effect size between HC and patients found in the large meta-analysis of the ENIGMA study group (Kelly et al., 2018) and similarly reported in our previous study (Pavan et al., 2025).

Future research on the WM microstructure alterations in SZ should explore their relationship with brain metabolism more directly. One potential approach is to examine the direct association between GSH-redox levels in the brain, measured through MR spectroscopy, and WM microstructure. Furthermore, exploring the involvement of other brain metabolites could also provide new insights into WM alterations.

## Limitations

Our study has a few limitations that need to be considered. First, we used harmonization procedures to remove both the effects of MRI scanner and of plate readers used to quantify the redox markers. In addition, the delay between MRI acquisition and blood collection could be another source of bias (Perkins et al., 2020) despite being capped and controlled during the statistical analyses. Second, the heterogeneity among the patient’s cohort (that includes early psychosis patients and subjects with chronic SZ) may significantly reduce the statistical power due to potential conflicting trends that may arise at WM microstructure or GSH-system levels across the different stages and types of psychosis. Third, we did not apply additional corrections across all modalities and contrasts for the cluster findings (e.g., a second false discovery rate pass). This decision stems from the large number of voxels resulting from pooling output images from multiple TBSS runs, and the inability of a second correction to account for correlations within the blood and microstructure measures. No finding will survive such conservative correction. Nevertheless, our clusters’ results, already corrected with the classical correction (Threshold-Free Cluster Enhancement + false discovery rate) found in TBSS, are large and spatially consistent, suggesting they are unlikely to be false positives. Fourth, lifestyle habits, such as smoking (Mons et al., 2016) and diet (Gould and Pazdro, 2019), could influence GSH, GPx and GR activities, having a confounding role on our results. Accounting for these factors in a reliable way is challenging as they are often self-reported. Lastly, GPx and GR activities and their ratio in blood cells inform us about the systemic redox homeostasis which does not necessarily reflect the conditions found in the brain. However, several studies have found associations of peripheral markers of the GSH-redox cycle with brain GSH levels (Xin et al., 2016), brain atrophy and enlargement of the lateral ventricles (Buckman et al., 1987), or EEG sensory evoked responses (Geiser et al., 2017).

## Conclusions

Altogether, our association analyses indicate that there is no direct correlation between the peripheral GSH-redox cycle and WM anomalies detected in PT, although a link between reductive stress and WM microstructure was found in HC. Our results do not therefore exclude the involvement of the GSH redox cycle during the development/maturation of WM. The dysregulation of this system during brain development could still contribute to WM anomalies in psychosis and schizophrenia.

## Acknowledgments

This work was supported by the Swiss National Science Foundation (PCEFP2_194260, to I.J.; 320030_197787 to PH and YA), the National Center of Competence in Research (NCCR) “SYNAPSY - The Synaptic Bases of Mental Diseases” from the Swiss National Science Foundation (n° 51NF40 – 185897 to KQD & PC) and the Foundation Alamaya. Dr. Alameda is supported by Carigest fellowship and by Frutiger Adrian et Simone fellowship. Dr. Dwir and P. Klauser are supported by Frutiger Adrian & Simone fellowship. We are grateful to Gloria Reuteler, Adeline Cottier and Morgane Baumgartner for expert technical assistance.

## Data Availability

Data will not be publicly available due to absence of consent from the participants. The code is available here: github.com/Mic-map/WMTI-Watson_Python

## Author Contributions

The authors confirm contribution to the paper as follows: study conceptualization: T.P, P.S., I.J, P.K., D.D.; investigation and formal analysis: T.P.; interpretation of results: T.P, P.S., I.J, P.K., D.D., R.J., Z.S.; writing — original draft: T.P., P.S, I.J.; MR data collection: M.C.; writing — review and editing: L.A., K.D., P.C., and P.H.

## Declaration of generative AI and AI-assisted technologies in the writing process

During the preparation of this work the author(s) used ChatGPT to improve language and readability. After using this tool/service, the author(s) reviewed and edited the content as needed and take(s) full responsibility for the content of the publication.

