## Supplementary Material for "Brain White Matter Microstructure Associations with Blood Markers of the GSH Redox cycle in Schizophrenia"

June 5, 2025

### Supplementary Material

### Methods

#### MRI Acquisition details

Magnetization-prepared rapid acquisition gradient echo (MPRAGE): echo time (TE) 2.98ms, repetition time (TR) 2300 ms, inversion time (TI) 900 ms, field of view (FOV) 160 x 240 x 256 $mm^3$ and voxel size 1 x 1 x 1.2 $mm^3$), acquisition time 7 minutes. Pulsed Gradient Spin-echo (PGSE) echo planar imaging (EPI) sequence: acquired in cartesian q-space coverage totalling 129 (Prisma) or 257 (Trio) DWI volumes. Prisma: TE = 144 ms, TR = 6.1 s; Trio: TE = 103 ms, TR = 5.9 s. FOV = 211 x 211 x 114$mm^3$, voxel size = 2.2 x 2.2 x 3$mm^3$, 96x96x38 slices, partial Fourier = 0.75, 1 b0 acquisition 128 (Prisma) and 256 (Trio) directions, acquisition time 13 minutes. Any participants with pacemaker, cochlear implant, implant of stimulator or drug pump, glucose sensor, bypass valve or pregnant were not allowed in the MRI scanner and excluded from the study.

#### MRI Preprocessing details

The diffusion preprocessing pipeline included MP-PCA denoising (Veraart et al., 2016) and Gibbs ringing correction (Kellner et al., 2016; Lee et al., 2021). The EPI distortions were corrected using ANTs non-linear registration since no reverse phase encode image or magnetic field map were acquired. The registration of the DSI b=0 volume to the MPRAGE was constrained only in the direction of the distortion, for then warping the estimated correction back to each DWI volume in native space (Alemán-Gómez et al., 2023; Tax et al., 2022). Thereafter, the distortion-corrected images were further corrected for eddy currents and motion using FSL *eddy* (Andersson and Sotiropoulos, 2016). Since FSL eddy does not support DSI data natively (see *–data_is_shelled* in: https://fsl.fmrib.ox.ac.uk/fsl/fslwiki/eddy/UsersGuide), temporarily merging or directly dropping a subset of volumes based on their b-values in order to simulate shells was necessary to accommodate the algorithm. Namely, volumes b=1500 and b=2000 were merged as b=1750, b=4000 was merged to b=4500 while b=6000 and b=8000 were removed. Right after the eddy execution the merged volumes were split back into their original b-values except for the dropped volumes (for the dMRI metric estimations only the b-values≤2500 were needed). The reduced DWI images featured 29 and 57 directions (Prisma, 1:b=0, 3:b=500, 6:b=1000, 4:b=1500, 3:b=2000, 12:b=2500; Trio, 1:b=0, 6:b=500, 12:b=1000, 8:b1500, 6:b=2000, 24:b=2500). Data quality was assessed by visual inspection and automated means (FSL's eddy QUAD and SQUAD (Bastiani et al., 2019)). The estimated quality metrics were volume-to-volume (absolute movement) and within-volume motion (relative movement), eddy current-induced distortions and data outliers. For the Trio scanner the study mean absolute movement was $1.08 \pm 0.39$ mm [min:0.21, max:3.21]; while the mean relative movement was $0.02 \pm 0.013$ mm [min:0, max:0.1]. For the Prisma scanner the study mean absolute movement was $0.46 \pm 0.31$ mm [min:0.13, max:3.36]; while the mean relative movement was $0.1 \pm 0.06$ mm [min:0.02, max:0.61]. Biophysical model metrics were inspected for outliers and voxels excluded across all the parametric maps if values were outside these biologically plausible ranges: $0 < FA < 1$, $0 < MD < 4$ $\mu m^2/ms$, $0 < MK < 10$, $0 < f < 1$. No other outlier condition was retained on the significant TBSS clusters that would lead to excluding subjects altogether from any given regression analysis.

### Blood sample quantification details

GR activity was assessed in hemolyzed blood cells incubated in phosphate buffer solution (100mM, pH 7.5) containing EDTA (0.6mM), and non-limiting levels of oxidized glutathione (GSSG, 2.5mM) and NADPH (0.25mM). The activity of GR was expressed as the amount of NADPH used by GR (in nmole/min at 22°C) to reduce GSSG. The decrease of NADPH was measured using the decrease of absorption at 340 nm (per min) and quantified using the NADPH 340 nm absorption coefficient. GPx activity was assessed in hemolyzed blood cells incubated in a phosphate buffer solution (100mM, pH 7.5) with EDTA (0.6mM), and non-limiting amounts of GSH (2.5mM), NADPH (0.25mM), GR (0.84U/ml; Fluka) and tert-butyl hydroperoxide (TBHP, 0.8mM, Fluka). The activity of GPx is proportional to the amount of NADPH used to reduce the GSSG produced during the reduction of TBHP by GPx. The decrease of NADPH was measured using the decrease of absorption at 340 nm (per min) and quantified using the NADPH 340nm absorption coefficient.

### Reader harmonization

To correct for the batch effect of the two microplate reader, the ComBat method from the R (R Core Team, 2023) package Surrogate Variable Analysis (SVA, Leek et al. (2012)) was used. GPx and GR values were harmonized together and we specified age, sex and clinical group as model matrix for outcome of interest so that the effect of the specified variables could be protected from accidental corrections. To assure the correction was working we compared the results obtained to the original data and the data normalized by reader type (demeaned and scaled by the standard deviation, see Fig. S 1 and 2). We observed that, despite the normalization already accounts for a large amount of the differences between the readers, the harmonization procedure better preserves the the relation within the reader group (in Fig. S 1 and 2, subplots G vs H).

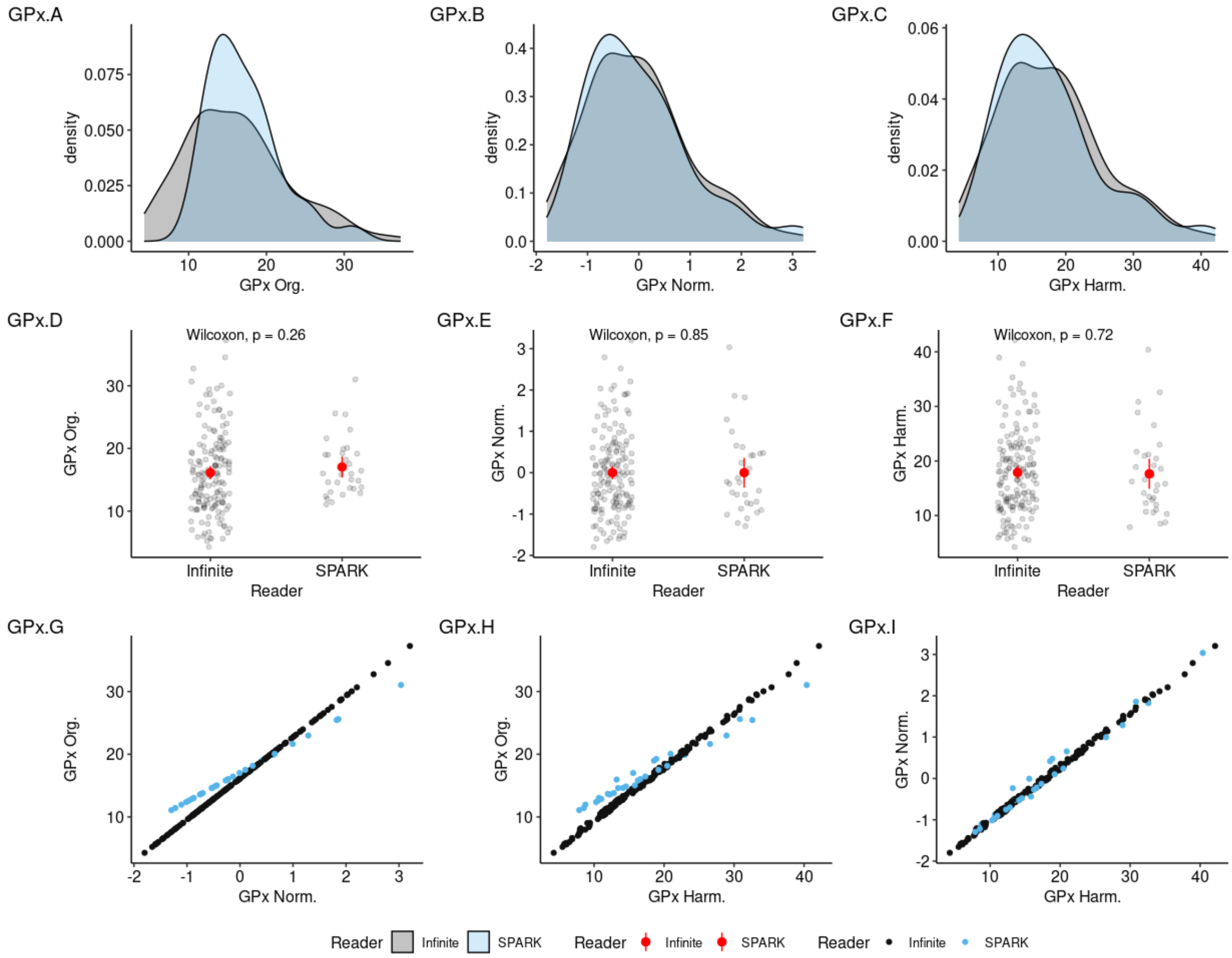


Figure S 1: comparison between GPx original data, normalization and harmonization. GPx distributions for original data (A), normalized by scanner (B) and harmonized (C). Group comparison between the readers of original data (D), normalized by scanner (E) and harmonized (F). Relation between original and normalized GPx data (G), between original and harmonized GPx data (H), between normalized and harmonized GPx data (I). Org.: Original; Norm.: normalised; Harm.: harmonized.

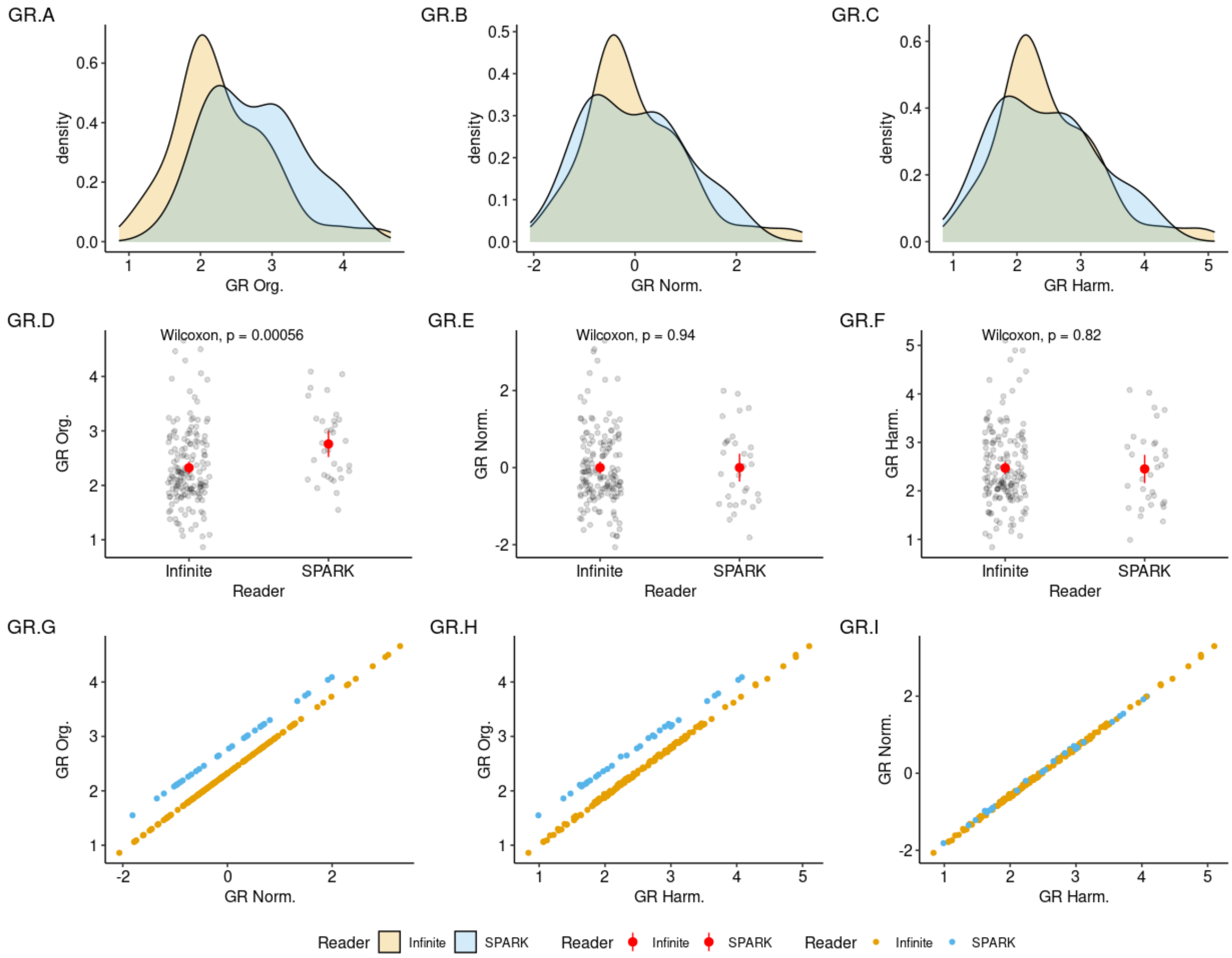


Figure S 2: comparison between GR original data, normalization and harmonization. GR distributions for original data (A), normalized by scanner (B) and harmonized (C). Group comparison between the readers of original data (D), normalized by scanner (E) and harmonized (F). Relation between original and normalized GR data (G), between original and harmonized GR data (H), between normalized and harmonized GR data (I). Org.: Original; Norm.: normalised; Harm.: harmonized.

### Age correction

To better model age and correct for its effect, five different model were evaluated for each of the 4 WM skeletons metric: no age effect (intercept-only), linear age effect (LAE), quadratic age effect (QAE), linear age effects with group interaction (LAEG) and quadratic age effects with group interaction (QAEG); for a total of 20 models. The model selection criteria was to minimize the Bayesian information criterion (BIC) between the fitted model for one metric, and then selecting the most popular correction model among the metrics. The procedure returned the QAE as the best model. See Figure S3 for an example.

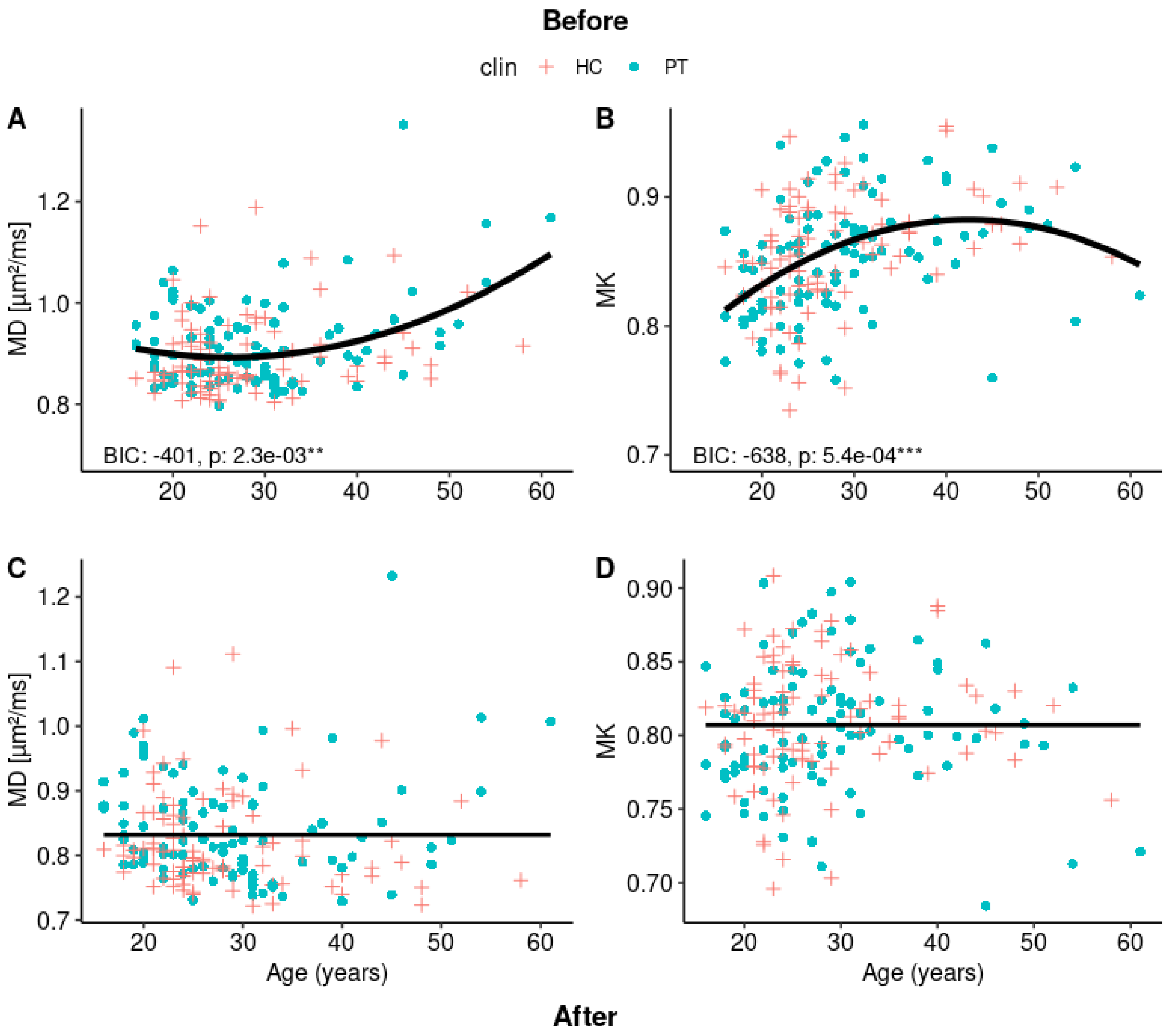


Figure S 3: Before (A, B) vs after (C, D) examples of fits for the best age correction model of the WM skeleton MD (A, C) and MK (B, D). The procedure selected the quadratic age effect (QAE) as best model. In the before-plots (A, B), the black solid line indicates the predicted values of MD or MK given the corresponding age. In the after age correction plots (C, D), the model intercept is maintained and just the age effect is removed.

### Diagnosis

| | | N | Pct. |
|---|---|---|---|
| Diagnosis | Schizoaffective disorder | 8 | 9.8 |
| | Schizophrenia | 65 | 79.3 |
| | Schizophreniform disorder | 9 | 11.0 |

Table S 1: Patients individual diagnosis. Pct. Percentage

### Delay

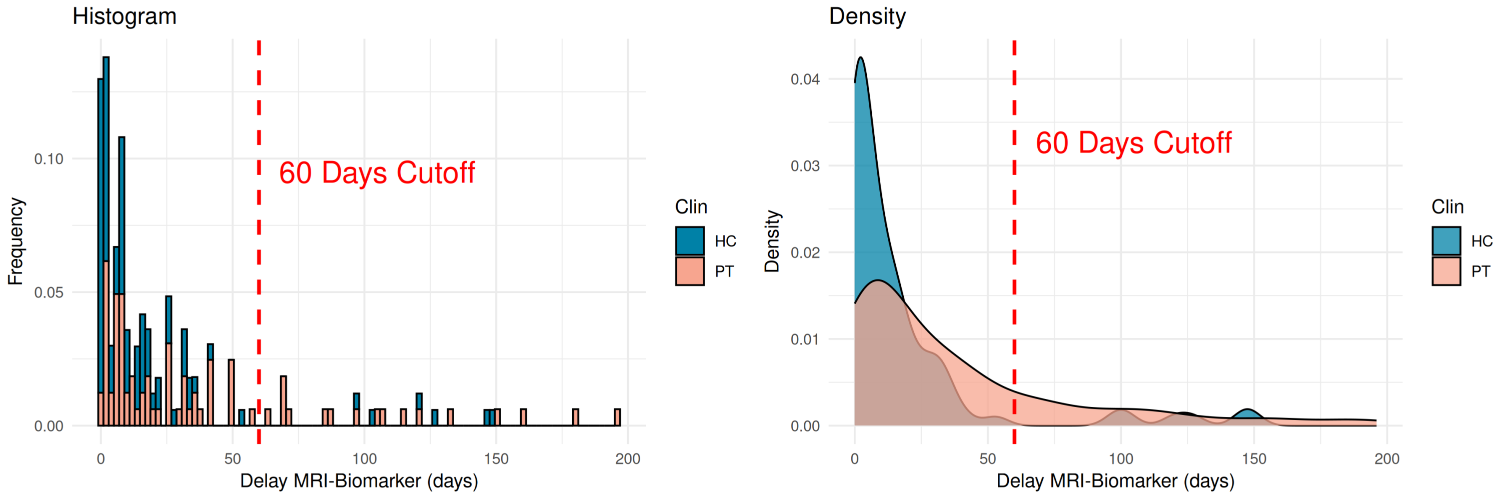


Figure S 4: Histogram and density plot of the absolute delay between blood biomarkers sampling and MRI scan. We found a significant difference in average delays between PT and HC (p=0.000044), and a non-negligible group of PT and HC participants had a large delay between blood sampling and MRI scan. In addition, blood GPx and GR measures decrease over several years with aging (Martínez De Toda et al., 2019; Jones et al., 2002). Thus, we applied a threshold of 60 days to the delay variable to reduce the group-level delay effect. It is unlikely that the effect of age-related variation, or other large fluctuations, could influence the GPx or GR markers within a 60 day window (Garcia et al., 2000). The 60 day threshold excluded the long tails (and heavier tail in PT) of the delay distribution, which reduced the delay difference between PT and HC ($p_{before}$=0.000044 to $p_{after}$=0.0012) without sacrificing a large number of participants (86% of the cohort was retained). However, the difference remained significant, so we included delay as a regressor to control for potential confounding effects.

### Associations between blood markers and clinical tests

| Score | Marker | Estimate | p-value |
|---|---|---|---|
| GAF | GPx | -0.16 | 0.66 |
| PANSS: Pos. | GPx | 0.017 | 0.88 |
| PANSS: Neg. | GPx | 0.17 | 0.23 |
| PANSS: Gen. | GPx | -0.16 | 0.55 |
| GAF | GR | 3.71 | 0.042 * |
| PANSS: Pos. | GR | -0.36 | 0.67 |
| PANSS: Neg. | GR | -0.04 | 0.97 |
| PANSS: Gen. | GR | -1.95 | 0.25 |
| GAF | GPx/GR-ratio | -0.95 | 0.29 |
| PANSS: Pos. | GPx/GR-ratio | 0.028 | 0.81 |
| PANSS: Neg. | GPx/GR-ratio | 0.49 | 0.14 |
| PANSS: Gen. | GPx/GR-ratio | 0.023 | 0.97 |

Table S 2: Correlation between redox blood biomarkers and clinical scores in patients. Note, the significant association do not survive multiple comparison correction. GPx: glutathione peroxidase, GR: glutathione reductase GAF: Global Assessment of Functioning, PANSS POS: PANSS Positive Subscale, PANSS NEG: PANSS Negative Subscale, PANSS GEN: PANSS General Psychopathology Subscale.
